# Controlling long-term SARS-CoV-2 infections is important for slowing viral evolution

**DOI:** 10.1101/2021.04.10.21255251

**Authors:** Debra Van Egeren, Alexander Novokhodko, Madison Stoddard, Uyen Tran, Diane Joseph-McCarthy, Arijit Chakravarty

## Abstract

The rapid emergence and expansion of novel SARS-CoV-2 variants is an unpleasant surprise that threatens our ability to achieve herd immunity for COVID-19. These fitter SARS-CoV-2 variants often harbor multiple point mutations, conferring one or more traits that provide an evolutionary advantage, such as increased transmissibility, immune evasion and longer infection duration. In a number of cases, variant emergence has been linked to long-term infections in individuals who were either immunocompromised or treated with convalescent plasma. In this paper, we explore the mechanism by which fitter variants of SARS-CoV-2 arise during long-term infections using a mathematical model of viral evolution and identify means by which this evolution can be slowed. While viral load and infection duration play a strong role in favoring the emergence of such variants, the overall probability of emergence and subsequent transmission from any given infection is low, suggesting that viral variant emergence and establishment is a product of random chance. To the extent that luck plays a role in favoring the emergence of novel viral variants with an evolutionary advantage, targeting these low-probability random events might allow us to tip the balance of fortune away from these advantageous variants and prevent them from being established in the population.

## INTRODUCTION

The widespread deployment of biomedical interventions against SARS-CoV-2 has highlighted viral evolution as a significant potential risk in bringing the ongoing pandemic to an end. SARS-CoV-2 has a high mutational rate, similar to other RNA viruses, and a high mutational tolerance in key proteins, such as Spike, the molecular target for many biomedical interventions against the disease^1–3^. The emergence and expansion of novel SARS-CoV-2 variants over the past few months threatens to undermine the promise of a return to normalcy as a result of the newly deployed vaccines. In fact, many of these new variants are more capable of infecting cells, spreading between hosts, and/or evading natural immunity or therapeutics^4–6^. On a population level, natural selection has acted to rapidly increase the frequency of these more-fit SARS-CoV-2 variants, leading them to dominate the local viral population after emergence (Table S1).The emergence of fitter variants has also led in a number of cases to more severe disease outbreaks, and poses the threat of eventual reductions in vaccine efficacy^4,5,7,8^.

In addition to acting at the population level, natural selection has been shown to select for advantageous viral variants that are generated within individual patients infected with SARS-CoV-2. Longitudinal sequencing of SARS-CoV-2 from individual patients has revealed selection for multiple antibody-evading mutations, particularly in patients with long-term infections treated with convalescent plasma^9^. Additionally, studies have shown that individuals with impaired immune function can shed high levels of virus for weeks, creating an environment where SARS-CoV-2 is exposed to prolonged selection pressures favoring variants that can escape the immune response and/or are resistant to treatment^10–13^.

Understanding the factors driving the evolutionary process for SARS-CoV-2 could potentially allow us to design and use biomedical interventions in a way that hinders viral evolution, giving us the upper hand in managing the pandemic. In particular, there are several steps in the process of generating new advantageous variants that occur largely by random chance.

First, sufficient genetic diversity must be created within infected individuals through stochastic events. SARS-CoV-2 mutations are initially generated by random errors in viral replication within individuals with COVID-19. Deep sequencing studies have revealed that the SARS-CoV-2 viral population exists within the host as a quasispecies^14,15^, a population structure with a large number of related sequences arising from *de novo* mutations that occur during the course of infection^16,17^. Quasispecies genetic diversity has been shown to vary over time^15,18^, and deep sequencing studies have demonstrated a role for genetic drift^19^ and intrahost transmission bottlenecks^14^ as the virus moves from one region of the body to another. While genetic diversity provides opportunities for advantageous mutations to arise and expand due to natural selection within individuals infected with SARS-CoV-2, genetic drift may provide a barrier to the deterministic expansion of advantageous viral mutations at lower population sizes.

Next, viral variants generated within a COVID-19 patient are transmitted to new hosts. During this process, further stochasticity is introduced by the low numbers of viral particles required to start an infection in a new host, which creates a narrow transmission bottleneck^16,17^. Consistent with this bottleneck, sequencing studies have provided conflicting results with respect to the ability of intrahost variants to transmit and establish new SARS-CoV-2 lineages that have the potential to transmit widely. While a number of studies have shown this to be the case^16,17,20,21^, other studies have not been able to demonstrate transmission of viral lineages derived from intrahost evolution^11,22,23^. The stochasticity in inter-host transmission of specific viral lineages is compounded by the overdispersed nature of SARS-CoV-2 spread, where most onward transmission originates from a small number of individuals. Recent work suggests an important role for stochastic extinction of variants, as at least five new infections are required in order for a newly emergent variant to establish itself in the population^24^.

These stochastic factors in the evolution of SARS-CoV-2 represent a potential weakness that can be exploited in the design of intervention strategies to slow viral evolution. To the extent that the stochastic contribution of drift can be increased, the deterministic contribution of natural selection to the improvement of viral fitness can be weakened. In this study, we have used evolutionary dynamics to better understand the process by which fitter SARS-CoV-2 variants arise during infections and to identify means by which viral evolution can be slowed.

## RESULTS

### Single mutants that are fitter within patients are more likely to be transmitted to new hosts

To investigate mutation and selection dynamics of SARS-CoV-2 within hosts, we simulated stochastic viral evolution using a modified Wright-Fisher model (Fig. 1A, Methods). During each 12-hour replication cycle, virions from the previous generation are randomly selected with probability proportional to their fitnesses to replicate and produce a burst of *N*_b_ new viral particles in the next generation. Each replicating virion has a constant probability of generating a new single point mutation that is passed to all of its *N*_b_ progeny. The total number of virions present in each generation is given by estimates from sputum RT-PCR measurements^25^ (Fig. 1B, Methods). Unless stated, the parameter values for the simulations are those given in Table 1.

**Table 1.** Parameter values used for intrahost SARS-CoV-2 evolutionary dynamics simulations.

| Name | Description | Value | Source |
| --- | --- | --- | --- |
| $t_g$ | SARS-CoV-2 replication cycle length | 12 hours | <sup>49</sup> |
| $N_b$ | Viral burst size- number of virions produced by each infected cell | 1000 | <sup>49</sup> |
| $\mu$ | Within-host SARS-CoV-2 mutation rate (per replication cycle, per site) | $10^{-5}$ | <sup>49</sup> |
| $N_{trans}$ | Number of virions transmitted to new host | 10 | <sup>17</sup> |
| $R_0$ | Reproductive number- average number of new hosts infected by a single infected host | 1.5 | Similar to the transmission advantage measured for B.1.1.7<sup>5</sup> |
| $\sigma$ | Number of new infected hosts (per day) | 50,000 | Similar to recently reported numbers for the US |
| $L_{LT}$ | Mean (± standard deviation) of infection length in long-term SARS-CoV-2 shedders | 93.75 days ± 11.48 | <sup>48</sup> |

**Figure 1.**
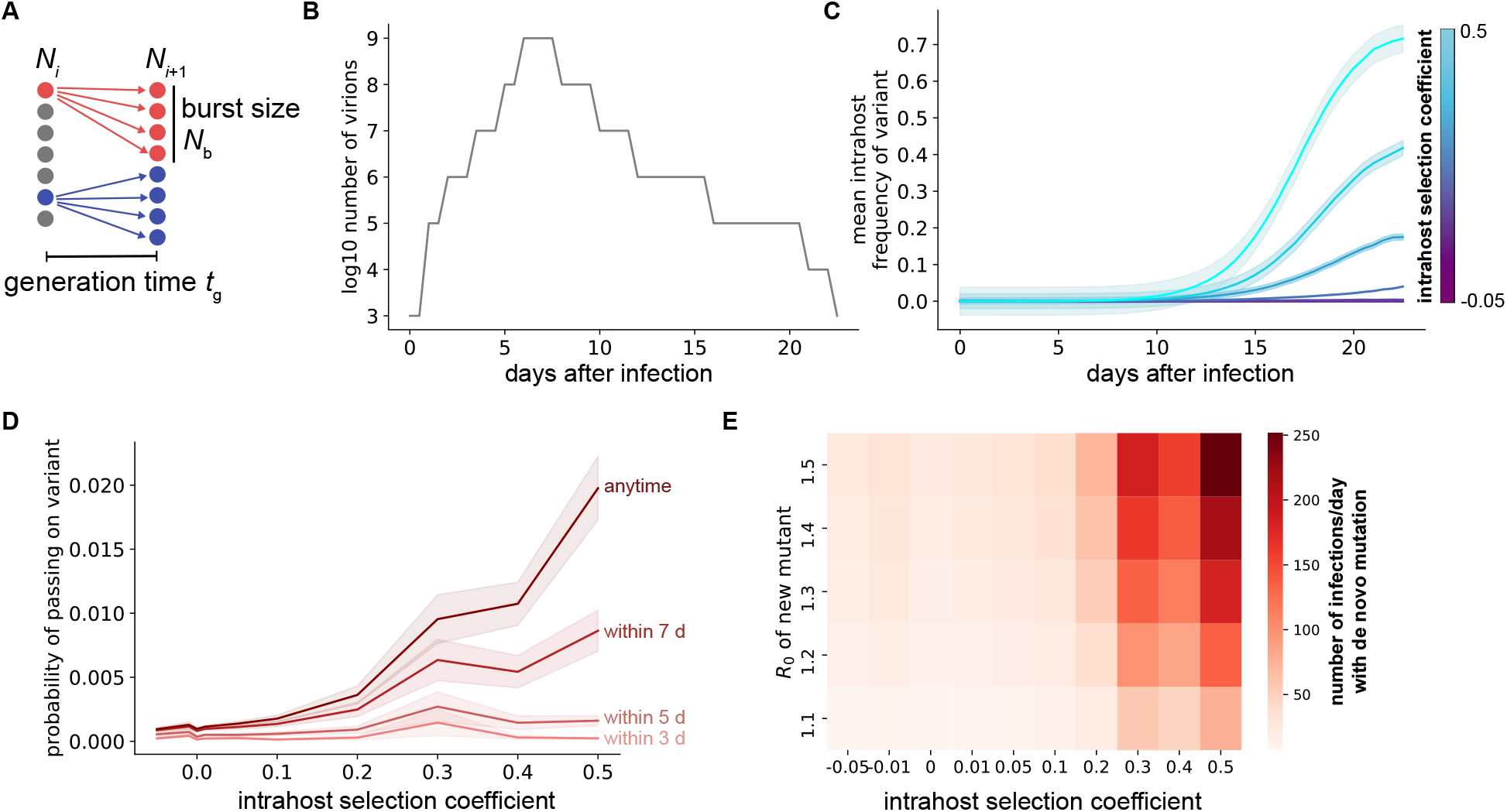
Selection within individuals with COVID-19 leads to selection of more fit viral variants. **A**. Schematic of viral replication model used to simulate SARS-CoV-2 evolutionary dynamics. **B**. Sputum viral load curve for a typical COVID-19 infection. The x-axis represents the time starting from the initial transmission event that started the infection. **C**. Mean frequency of variants with point mutations within individuals with COVID-19 for different mutation fitness effects (colors). **D**. Probability of a specific single mutation to be present in at least one virion transmitted if transmission occurs within the first 3-7 days of infection (lighter red curves) or anytime during infection (darker red). For **C** and **D**, shaded areas represent ± SEM, n=1000 simulations per condition. **E**. Number of total new single mutant infections generated per day that establish a surviving variant lineage, assuming all infections are of standard length with viral load profile given in **B** and that transmission occurs within the first 7 days of infection. Unless otherwise specified, simulation parameter values are those given in Table 1.

Over the course of a typical-length COVID-19 infection (23 days), our simulations show that viral variants with point mutations that increase the replication probability by 20-50% (selection coefficients of 0.2-0.5) expand significantly more than variants with neutral or weakly deleterious fitness effects (Fig. 1C). This expansion of fitter variants increases the probability that at least one viral particle with a specific beneficial mutation will be transmitted to a new host (Fig. 1D, Methods). Individuals are more likely to pass on beneficial variants if transmission occurs later in the infection, since the frequency of these variants increases over time due to selection. Assuming these variants are also able to spread through the population with a moderate transmission advantage, new lineages with advantageous single mutations are rapidly created at the population level (Fig. 1E). These results suggest that selection for beneficial single point mutations within COVID-19 patients increases the rate at which fitter SARS-CoV-2 lineages establish at a population level.

### Longer infection duration and higher viral load increase probability of transmitting fitter variants

Viral load dynamics vary significantly between COVID-19 patients. To assess the evolutionary consequences of this variation, we simulated viral replication and transmission of variants for patients with different viral load kinetics (Fig. 2). Patients with longer periods of peak viral load were able to transmit fitter SARS-CoV-2 variants more efficiently (Fig. 2A). On the other hand, decreasing viral load decreased the probability that fitter variants generated within a patient would be transmitted (Fig. 2B). Both of these observations indicate that the strength of selection increases as the number of replicating viruses over the course of an infection increases.

**Figure 2.**
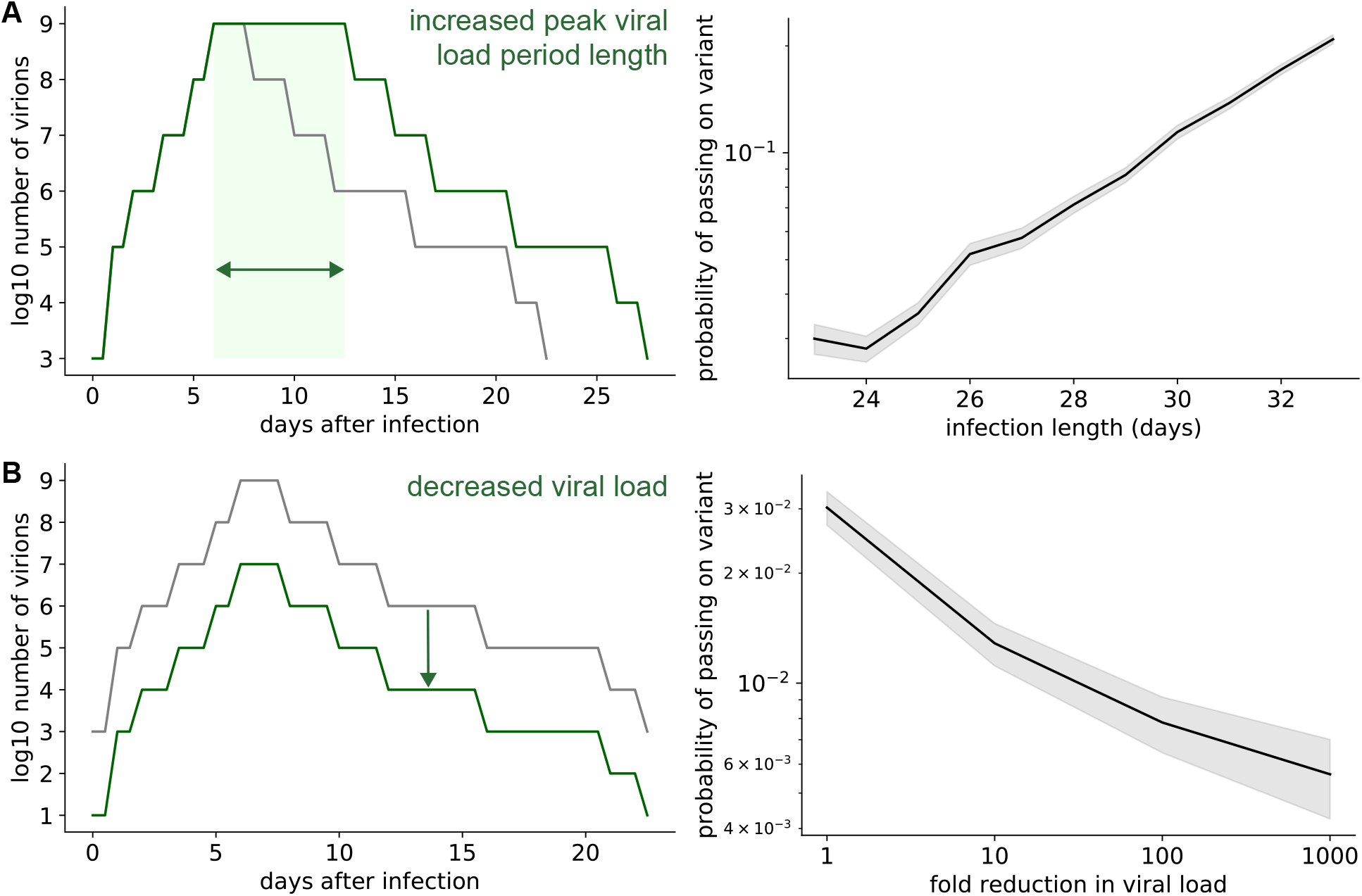
Within-host selection is stronger for infections that last longer or have higher viral loads. **A**. Probability that at least one transmitted virion has a specific advantageous single mutation, for different overall lengths of infection. Infection lengths were adjusted by increasing the length of the peak viral load period (left panel schematic). **B**. Probability that at least one transmitted virion has a specific single mutation, for different viral loads. Viral loads were adjusted by reducing the viral load by a constant factor over the entire course of infection (left panel schematic). For right panels, shaded areas represent ± SEM, n=1000 simulations per condition, with a selection coefficient of 0.2 for the single mutant. Unless otherwise specified, simulation parameter values are those given in Table 1.

### Beneficial two-mutation combinations are readily generated within patients with long-term SARS-CoV-2 infections

Many of the reported new SARS-CoV-2 variants are defined by more than one point mutation. One of these variants that arose during long-term SARS-CoV-2 infection within an immunocompromised individual included a mutation that reduced infectivity but conferred resistance to neutralizing antibodies, which was offset by a mutation that increased infectivity^9^. These observations suggest that highly-fit mutation combinations that require transit through a deleterious intermediate state (i.e., crossing a fitness valley) may be generated within COVID-19 patients with longer infection durations.

To investigate the rate at which these variants are generated within hosts, we modeled a multistep mutation process where beneficial mutation combinations are created from deleterious intermediates that only have some of the mutations found in the beneficial combination (Fig. 3A). Beneficial two-mutation combinations exist at very low frequencies over the timescale of a typical-length SARS-CoV-2 infection, but increase in frequency within hosts that have prolonged infections (Fig. 3B). This increase in variant frequency that occurs within hosts due to selection corresponds to an increase in the probability the beneficial two-mutation variant will be transmitted (Fig. 3C). Therefore, patients with longer SARS-CoV-2 infections are more likely to generate a variant with multiple mutations. On a population level, these individuals who produce and shed virus for long periods (> 30 days after symptom onset) increase the production rate of new variants with multiple mutations. Increasing the proportion of COVID-19 patients with prolonged SARS-CoV-2 shedding increases the rate at which new, fitter variants with two mutations are produced (Fig. 3D).

**Figure 3.**
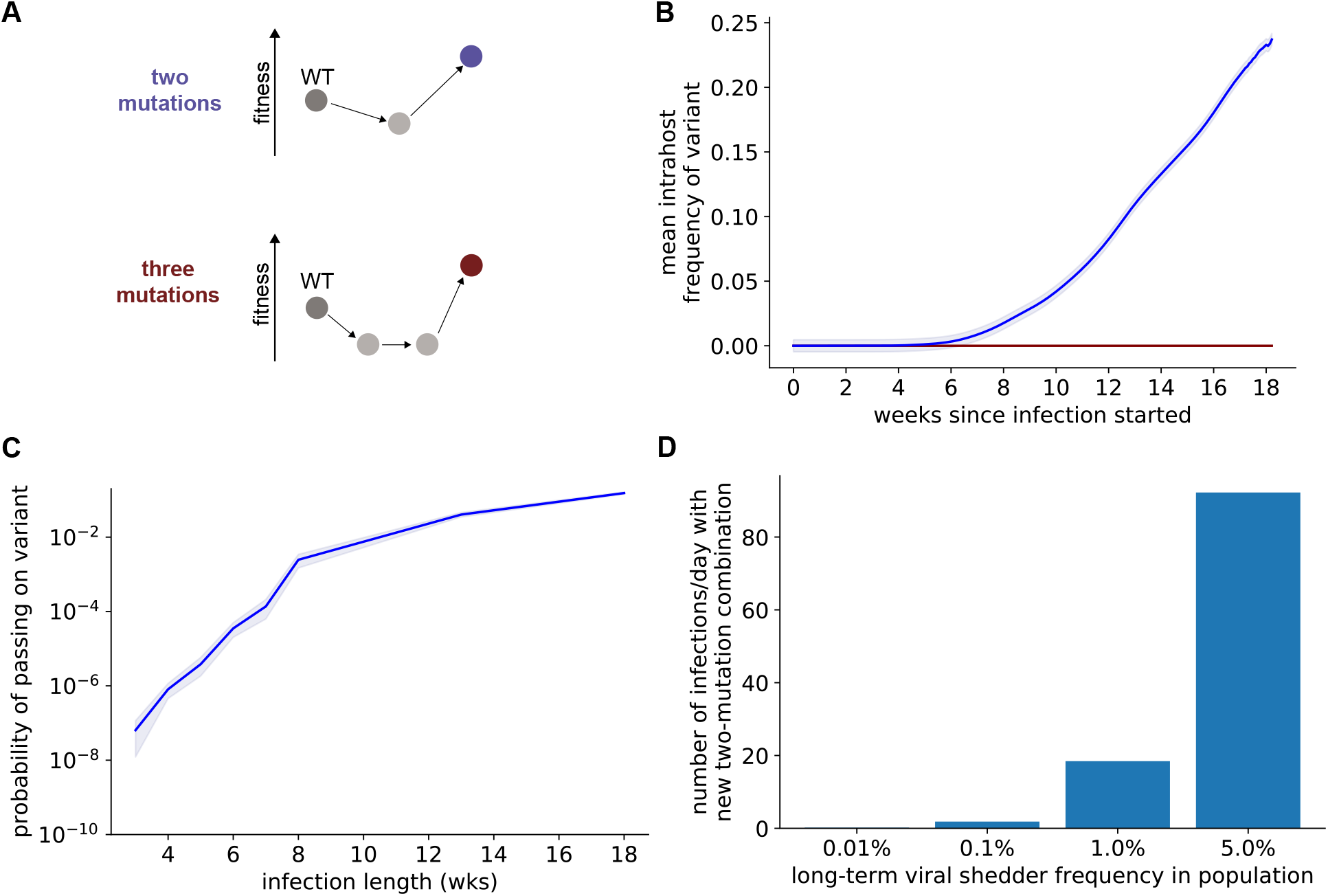
SARS-CoV-2 can acquire multiple mutations during infections with sustained viral replication. **A**. Fitness valley crossing model for acquisition of multiple mutations. Intermediate states with fewer mutations (light grey) have lower fitness than the WT virus within individuals, while variants with a specific combination of two or three mutations (blue and red, respectively) have higher fitness. **B**. Mean frequency of variants with a beneficial combination of two (blue) or three (red) mutations within individuals with long-term SARS-CoV-2 infection. **C**. Probability of a beneficial combination of two mutations (blue line) to be present in at least one virion transmitted if transmission occurs anytime during infection. For **B** and **C**, shaded areas represent ± SEM, n=1000 simulations per condition. **D**. Number of *de novo* double mutant infections that establish a surviving lineage when some COVID-19 patients shed live virus for more than 30 days after developing symptoms. Deleterious intermediates had a fitness cost of 0.05 and beneficial mutation combinations had a selective advantage of 0.2. Unless otherwise specified, simulation parameter values are those given in Table 1.

### SARS-CoV-2 evolution can be impeded by targeting stochastic events required for the emergence of new variants

Several steps are required for a new SARS-CoV-2 variant to be generated and established in the population (Fig. 4A), and each of these steps represents a potential choke point for viral evolution that can be exploited in the design of interventions. First, a new variant must be generated through mutation and expand within a host. The efficiency of generating and selecting advantageous variants within COVID-19 patients can be reduced by biomedical interventions that reduce viral load within patients or reduce the frequency of long-term infections which lead to viral transmission (Fig. 4B). Variants then must be transmitted to additional hosts to establish within the population. Reducing the number of viral particles transmitted to new hosts (for example, through mask wearing or vaccination) will also slow the rate of emergence of advantageous variants (Fig. 4B). Finally, interventions specifically aimed at reducing the transmissibility of the new variant would also reduce their probability of spreading widely (Fig. 4B). This could be achieved by deploying a patchwork of vaccines and other prophylactics that target distinct epitopes, deliberately increasing the diversity of biomedical interventions to provide a more challenging evasion landscape for the virus. Taken together, our simulation results suggest that there are several low-probability stochastic events that are important for SARS-CoV-2 variant emergence and that interventions targeting these events can slow SARS-CoV-2 evolution.

**Figure 4.**
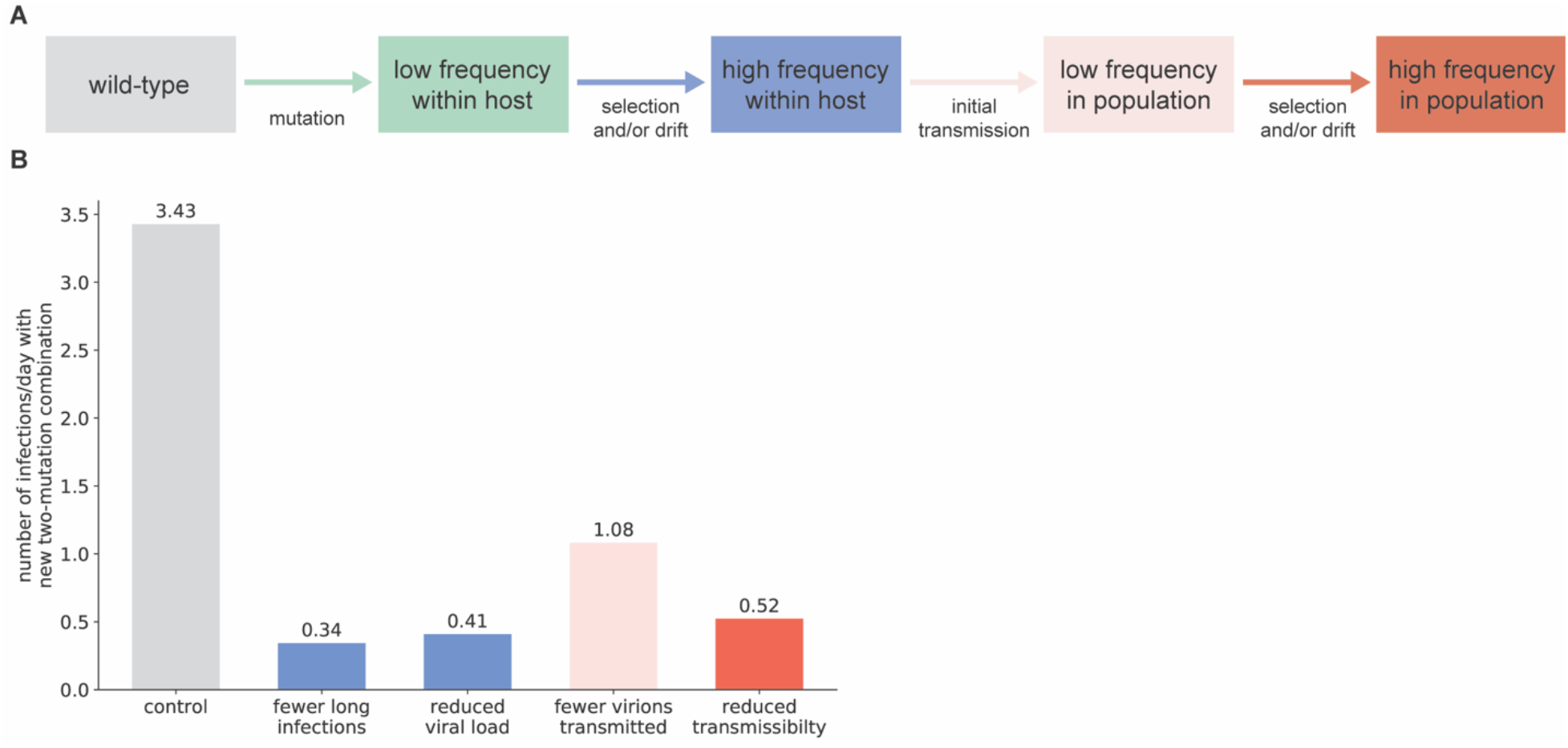
Interventions targeting stochastic events during infection or transmission reduce the efficiency of SARS-CoV-2 evolution. **A**. Schematic showing events necessary for the generation and population-level establishment of a new SARS-CoV-2 variant. **B**. Reduction in generation rate of new double mutant lineages that establish in the entire population caused by interventions targeting different events required to create a new surviving viral variant (n=1000 simulations per condition). Each intervention reduces the stated parameter value by 90%. Parameter value changes are relative to the control parameter set given in Table 1, with 0.1% of new infections lasting >30 days after symptom onset. The “reduced transmissibility” condition refers to a 90% reduction in the transmission advantage of the variant, leading to an *R*_0_ of 1.05.

## DISCUSSION

The recent emergence of variants of SARS-CoV-2 with multiple mutations poses a direct threat to the viability of vaccine-based suppression strategies for the pandemic^26^. A number of phenotypic changes that are capable of providing a fitness advantage have been associated with newly emerged variants. These include increased transmissibility, lethality, and viral replication rate, all characteristics of the B.1.1.7 lineage^5,27,28^. Additionally, some variants have immune evading phenotypes, including increased reinfection potential (e.g., B.1.351)^29^, and reduced neutralization for monoclonal antibodies (e.g., B.1.1.7, P.2 and B.1.351)^30,31^, convalescent plasma (e.g., B.1.351, B.1.429)^29,32^, vaccine induced sera (e.g., B.1.351, B.1.429, B.1.1.7, B1.298, P.2, and P.1)^33,34^ or complete vaccine evasion (B.1.351)^34^ (see Supplemental Material for further details). Some viral variants have also been associated with a longer duration of infection^35^, which is particularly concerning in the light of our finding that longer infections increase the probability of transmitting fitter SARS-CoV-2 variants.

In this work, we investigated the impact of within-host SARS-CoV-2 mutation and selection on the emergence of new viral variants in the population. Using a stochastic computational model of viral replication and selection, we found that mutations that increased the rate of viral replication increased in frequency within hosts during the course of a typical SARS-CoV-2 infection. This expansion led to more frequent transmission of the new variant and faster emergence of the variant on a population level. The effect of selection within hosts is more pronounced for longer SARS-CoV-2 infections with higher viral load, which can lead to the generation of variants with multiple mutations within a single host during prolonged infection. Thus, our work provides a quantitative understanding of the evolutionary mechanisms by which prolonged SARS-CoV-2 infections lead to the emergence of fitter viral variants.

This work, along with other recent findings on the stochastic nature of SARS-CoV-2 transmission^24^, suggests strategies that can be used to suppress the emergence of these fitter viral variants, thereby slowing the evolution of SARS-CoV-2. First, long-term SARS-CoV-2 infections (lasting longer than 30 days) should be treated as a serious public health concern, regardless of the presence of symptoms. Our work suggests that the low frequency of such long-term cases belies the threat that they pose to public health. In particular, asymptomatic long-term infections still transmit^10–13^ and can potentially be efficient accelerators of viral evolution. Isolation measures for such long-term infections should be designed to minimize transmission in the interest of minimizing their disproportionate impact on the pace of viral evolution. This would represent a departure from current practice (the CDC, for example, does not require negative testing prior to ending isolation for mild and asymptomatic casis of COVID-19^36^). Second, treatments for long-term viral infections should be aimed at the suppression of viral load and should be initiated regardless of whether patients are symptomatic. Such treatment regimes should consist of at least three active agents that can suppress viral replication and preferably target viral proteins other than Spike to reduce the risk of generating viral mutants that are resistant to the treatment. This shift in medical practice may reduce the risk of intrahost evolution leading to emergence of immune-evading SARS-CoV-2 variants, as likely occurred in some documented cases with convalescent plasma treatment^9^. Finally, contact tracing of transmission events and genetic characterization of secondary infections resulting from long-term infections (or from immunosuppressed patients) will be crucial.

We note that detecting long-term infections with SARS-CoV-2 represents a major logistical challenge in its own right-a widely deployable method for distinguishing long-term infections from spurious PCR results is an unmet need at present. While viral culture can be used to confirm the presence of replication-competent virus^37^, logistical constraints (such as turnaround time and the requirement for Biosafety Level 3 containment) limit its clinical use^38^. Subgenomic RNA (sgRNA) has been proposed as a biomarker for actively replicating virus^39^, but this remains controversial at present^40^. Therefore, there is an urgent need to develop a validated biomarker for viral replication in long-term infections.

Our work also suggests reasons for concern if intrahost evolution in long-term SARS-CoV-2 infections is not addressed directly. As intrahost evolution leads to faster generation of beneficial variants at a population level, new waves of transmission driven by fitter viral variants can be expected to arise as a result of untreated long-term infections. Our results also suggest another evolutionary incentive favoring increased infection duration, since long-term SARS-CoV-2 infections increase the rate at which the virus evolves. To the extent that this evolution is shaped by immune evasion, this study also points out an evolutionary route that is open to SARS-CoV-2, by which the virus can evolve to become progressively more lethal. Notably, while this potential risk conflicts with recent work that has conjectured that SARS-CoV-2 evolution may lead to progressive decreases in viral virulence^41^, it is consistent with available real-world evidence for this virus so far^27^. The evolution of increased virulence over time has been observed for several other viral pandemics, including for HIV over the years 1984-2010^42^, the second wave of the 1918-1919 Influenza pandemic^43^, and myxomatosis in rabbits in the 1970s and 80s^44–46^.

The deployment of multiple vaccines against SARS-CoV-2 in under a year is a remarkable triumph of modern medicine. These vaccines hold the promise of bringing the current pandemic to an end, but are vulnerable to the rapid emergence and expansion of immune-evading viral variants. Understanding the mechanism of SARS-CoV-2 evolution allows us to design strategies that can tip the balance in this evolutionary arms race and ultimately allow us to control the spread of SARS-CoV-2.

## METHODS

### Intrahost viral dynamics simulations

We used a modified Wright-Fisher evolutionary model to investigate viral mutation and selection dynamics within individuals with COVID-19. This model assumes that SARS-CoV-2 virions replicate in discrete generations of length *t*_g_ and that each replicating virion produces a fixed burst size of new virions *N*_b_. The total number of virions present at each generation was estimated from nasopharyngeal swab and sputum qRT-PCR measurements after diagnosis^25^. During each replication cycle, *N*_t_/*N*_b_ virions from the previous generation are chosen to reproduce, where *N*_t_ is the total number of virions in the current generation and *N*_b_ is the burst size. Each virion from the previous generation has a probability of reproducing and contributing *N*_b_ virions to the next generation that is proportional to its fitness. During each replication, there is a constant probability of generating a particular mutation that will be present in all *N*_b_ virions that are produced by that parent virion. Parameter values used for simulations are given in Table 1.

### Probability of variant transmission from a single infected host

The probability that a transmission event that occurs within a particular time window during infection was estimated by assuming new infections are initiated by *N*_trans_ virions sampled uniformly at random from the transmission period which go on to establish a new infection. Therefore, the probability that at least one virion with a specific mutation is passed on is 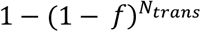, where *f* is the fraction of virions with the mutation that existed within the transmission window.

### Population-level de novo generation rate of SARS-CoV-2 variants

A newly-generated variant present in a single patient will initially spread stochastically to new hosts, the success of which will determine whether the variant will survive in the population. We modeled this stochastic spread as a branching process in which infected individuals infect on average *R*_0_ new hosts. More specifically, each infected individual transmits to *X* new hosts, where *X* is a random variable distributed as Poisson(*R*_0_). Probability theory results for Poisson branching processes states that if a new variant has *R*_0_ > 1, it will survive with probability *π*, where *π* is the smallest solution to 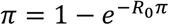 in the interval [0,1]^47^.

The rate at which new surviving variants are generated from patients with infections of length *i* is *πσ*_*i*_*p*_*i*_, where *p*_*i*_ is the probability of a patient with infection length *i* generating and transmitting the variant and *σ*_*i*_ is the number of patients with infection length *i* that are infected each day. The total rate at which new surviving variants are generated in the population is just the sum over all infection lengths, ∑_*I*_ πσ_*i*_*p*_*i*_. To estimate *σ*_*i*_, we assumed that most infections were of the typical length and viral load kinetics shown in Fig. 1B. However, a small fraction *p*_LT_ of infections were assumed to be longer than 30 days. The distribution of infection lengths in these long-term patients was assumed to be normal, and mean and standard deviation were estimated from longitudinal oral swab RT-PCR data^48^. The rate *z*_LT_ at which new surviving variants are generated from long-term patients was estimated by simulating infections that are 1, 2, 3, 4, 5, 10, or 15 weeks longer than typical infections to estimate *p*_*i*_ for each infection length and multiplying by the fraction of long-term infections within the length interval ending at *i*. That is, *z*_*LT*_ = ∑_*i*_ πσ_*i*_*p*_*i*_ for *i* in {30, 37, 44, 51, 58, 95, 128} days, where *σ*_*i*_ = P(*X* < *i*) -*σ*_*i*-1_ and *X* is normally distributed according to the distribution of infection lengths. The total rate at which new surviving variants are generated in the entire population is therefore *z*_LT_ + *z*_ST_, where *z*_ST_ is the rate at which standard length infections produce the variant.

## Data Availability

All simulation data are presented in the manuscript. No new experimental or clinical data were collected.

## ACKNOWLEDGEMENTS

None.

## AUTHOR CONTRIBUTIONS

DVE, MS, and AC conceived the original study design. DVE and AC designed the mathematical modeling approach and wrote the first draft. DVE conducted the modeling, with input from MS and AC. AN and UT contributed text to the Introduction, Discussion and Supplemental Materials. All authors edited and provided input into the manuscript.

## SUPPLEMENTAL MATERIAL

### Notes on phenotypic changes conferring fitness advantages in SARS-CoV-2 variants

Several variants with phenotypic changes have been characterized. Increased transmissibility (which has been observed *in vivo* with spike protein mutations such as D614G^50^) best explains the rapid spread of B.1.1.7 in multiple countries^5^. Greater lethality has been observed for the B.1.1.7 variant^27^ along with faster replication (e.g. B.1.1.7)^28^. Increased potential for reinfection is seen for B.1.351^29^. Multiple variants evade antibody therapies: for instance B.1.526, B.1.429, B.1.427, P.1 and B.1.351 are all resistant to spike-targeting monoclonal antibody bamlanivimab^51^. In other cases, reduced susceptibility to vaccines has been reported. In the extreme case, the variant B.1351 wholly evades the ChAdOx1 vaccine^34^. *In vitro* work found that sera from people vaccinated with CoronaVac does not neutralize the P.1 variant^52^. This explains the reported lower efficacy of CoronaVac in clinical trials in Brazil, where P.1 is prevalent, when compared to the United Arab Emirates. *In vivo* data, such as sequencing of breakthrough infections, would strengthen or falsify this hypothesis. Some viral variants have also been associated with longer duration of infection^35^, which is particularly concerning in the light of the findings in this report.

This list is not comprehensive, and new data are constantly being reported.

**Supplementary Table S1.** Emergence of currently common SARS-CoV-2 variants.

| **Variant** | **Site of Emergence** | **Date first reported** | **Local case frequency since emergence as of 4/4/2021** |
| --- | --- | --- | --- |
| B.1.351 | South Africa | October 2020<sup>53</sup> | 1668/2301 (72.5%)<sup>54</sup> |
| B.1.1.7 | UK | September 2020<sup>53</sup> | 173624/277811 (62.5%)<sup>54</sup> |
| P.1 | Brazil | January 2021<sup>55</sup> | 584/1519 (38.4%)<sup>54</sup> |

The B.1.351 variant was first identified in South Africa and quickly became the dominant variant in South Africa and in many other countries. As of April 4th 2021, the B.1.351 variant accounted for 72.5% of the case frequency in Brazil, and 7 other countries have at least a 72% frequency of B.1.351. This variant is known to increase the transmission rate and moderately impact neutralization by monoclonal antibodies^56^.

The B.1.1.7 variant first emerged in the UK in December 2020. By late December 2020 (week 52 of 2020), B.1.1.7 became the dominant variant, with a frequency of 51.31%. This variant is known to increase transmissibility and may increase risk of death compared with other variants^5,27^.

There are also some shared mutations among these three variants. The P.1. variant and B.1.351 variant share three mutations in the Spike protein, while the P.1. variant and B.1.1.7 share the N501Y mutation^56^.

